# Quantifying antibody kinetics and RNA shedding during early-phase SARS-CoV-2 infection

**DOI:** 10.1101/2020.05.15.20103275

**Authors:** Borremans Benny, Gamble Amandine, KC Prager, K Helman Sarah, M McClain Abby, Cox Caitlin, Savage Van, James O Lloyd-Smith

## Abstract

Our ability to understand and mitigate the spread of SARS-CoV-2 depends largely on antibody and viral RNA data that provide information about past exposure and shedding. Five months into the outbreak there is an impressive number of studies reporting antibody kinetics and RNA shedding dynamics, but extensive variation in detection assays, study group demographics, and laboratory protocols has presented a challenge for inferring the true biological patterns. Here, we combine existing data on SARS-CoV-2 IgG, IgM and RNA kinetics using a formal quantitative approach that enables integration of 3,214 data points from 516 individuals, published in 22 studies. This allows us to determine the mean values and distributions of IgG and IgM seroconversion times and titer kinetics, and to characterize how antibody and RNA detection probabilities change during the early phase of infection. We observe extensive variation in antibody response patterns and RNA detection patterns, explained by both individual heterogeneity and protocol differences such as targeted antigen and sample type. These results provide a robust reference for clinical management of individual patients, and a foundation for the mathematical models and serological surveys that underpin public health policies.

## Introduction

Since its emergence in December 2019, the SARS-CoV-2 pandemic has been the subject of intense research assessing all facets of the pathogen and its rapid global spread. Serology – the measurement of serum antibodies – provides crucial data for understanding key aspects of infection (Weitz *et al*. 2020). At the level of populations, serologic data can provide insights into the spread of the virus by enabling estimation of the overall attack rate, and seroprevalence estimates can elucidate the potential for herd immunity (Stringhini *et al*. 2020). In addition, these estimates are essential for developing accurate mathematical models of virus transmission dynamics, which provide the foundation for policies to reopen societies (German *et al*. 2020; Kissler *et al*. 2020; Krsak *et al*. 2020). At the level of individuals, the presence and concentration of antibodies against SARS-CoV-2 are indicators of past exposure, providing insights into incidence over a much wider temporal window than other metrics. When considered jointly with PCR testing to detect viral shedding, antibodies substantially improve the probability of detecting present and past infections (Prager *et al*. 2019), which is highly valuable given that RNA shedding is typically limited to a relatively brief infectious period, and because PCR sensitivity varies considerably with infection severity and biological sample type (Yongchen *et al*. 2020; Zhang *et al*. 2020b). Assessment of the levels of different antibody types (e.g. IgG, IgM) may even be used to infer approximately when individuals became infected (Chang *et al*. 2005; Borremans *et al*. 2016; Du *et al*. 2020), while detection of neutralizing antibodies may indicate protection from reinfection (Ni *et al*. 2020).

These applications of serologic data depend critically on knowing when different antibodies against the pathogen become detectable (seroconversion time), how their concentrations change over time (antibody level kinetics) and how long they last (antibody decay) (Lipsitch *et al*. 2020). When these key factors are known, serologic data become a powerful tool for inferring infection attack rate and transmission dynamics in the population (Winter & Hegde 2020). Five months into the pandemic, a remarkable number of serologic studies on the initial immune response against SARS-CoV-2 have been published. These studies were conducted in different laboratories, used different assays and sampling methods, and were performed on different patient groups that showed different manifestations of SARS-CoV-2 infection (Kontou *et al*. 2020; Lassaunière *et al*. 2020; Whitman *et al*. 2020).

This extensive variation arising from different sources creates a significant challenge for integrating existing data into one coherent picture of antibody kinetics and viral RNA shedding following SARS-CoV-2 infection. For example, in 23 studies reporting the kinetics of anti-SARS-CoV-2 antibodies, we found the use of 8 different antibody assays, 10 different target antigens, and 9 different reported antibody level units (studies are listed in the methods section). Additionally, the temporal resolution at which studies are conducted is highly variable: while some studies report antibody measurements for specific days, many group results into periods of multiple days or even weeks. Integrating such diverse results is therefore challenging, and requires statistical methods specifically developed for these data. This type of integration is however essential to capitalize on the limited and diverse data available to assess to what degree antibody and RNA shedding patterns are affected by assay type and target antigen choice. Properly integrated measures would also enable us to test whether disease severity and antibody patterns are linked, as results published to date are currently inconclusive (Huang *et al*. 2020; Tan *et al*. 2020).

In this study we quantified IgG and IgM antibody kinetics and RNA shedding probability during SARS-CoV-2 infection (up to 60 days post symptom onset) by drawing on published data. We formally characterized IgG and IgM seroconversion times, detection probabilities over time and antibody level kinetics using methods tailored to accommodate the diverse ways in which data have been collected and reported. We investigated how these variables are affected by disease severity, assay type and targeted antigen, and how patterns differ between IgG and IgM. We also assessed how antibody level kinetics relate to the probability of detecting viral RNA in various biological samples. For all variables we estimated mean values as well as observed variation in order to provide the complete picture required to interpret serological data, inform mitigation strategies and parameterize mathematical models of pathogen transmission while accounting for uncertainty. This formal integration approach enabled us to leverage 3,214 data points from 516 individuals with symptoms ranging from asymptomatic to critical, published in 22 studies, resulting in a quantitative synthesis of diverse data on anti-SARS-CoV-2 antibody patterns and RNA shedding during the early phase of infection.

## Methods

### Article selection

We considered preprints and peer-reviewed articles reporting the presence (positive or negative) or levels for IgG, IgM or neutralizing antibodies against SARS-CoV-2 or SARS-related CoV RaTG13 measured by enzyme-linked immunosorbent assay (ELISA), magnetic chemiluminescence enzyme immunoassay (MCLIA), lateral flow immunoassay (LFIA) or plaque reduction neutralization test (PRNT). In addition, we considered studies reporting PCR data. Only data associated with information about time since symptom onset at the moment of sample collection were included in the study. Different combinations of search terms were used in order to maximize the likelihood of finding an article through Pubmed, Google Scholar, and medRxiv. Articles (N = 23) available up to May 1 2020 containing data that can be used for the analyses in this study were included (Adams *et al*. 2020; Du *et al*. 2020; Thevarajan *et al*. 2020; To *et al*. 2020; Wölfel *et al*. 2020; Xiang *et al*. 2020; Xiao *et al*. 2020; Yongchen *et al*. 2020; Young *et al*. 2020; Zhang *et al*. 2020a, b, c; Haveri *et al*. 2020; Zhao *et al*. 2020; Zhou *et al*. 2020; Zou *et al*. 2020; Jiang *et al*. 2020; Lee *et al*. 2020; Liu *et al*. 2020a, b; Long *et al*. 2020; Lou *et al*. 2020; Okba *et al*. 2020). Data were extracted from published material, and digitized from figures when necessary using WebPlotDigitizer (Rohatgi 2019).

### Disease severity classification

Disease severity information was classified into three groups: asymptomatic/subclinical, mild/moderate, and severe/critical. Individuals were assigned a classification of asymptomatic/subclinical (N = 11) if they were referred to as “healthy”, “having no symptoms related to COVID-19”, or “asymptomatic”. Inclusion criteria for classification as mild/moderate or severe/critical are based on definitions from the Centers for Disease Control and Prevention (CDC)^1^, the Chinese National Health Commission (National Health Commission & State Administration of Traditional Chinese Medicine 2020), and the World Health Organization (World Health Organization 2020). When disease severity was not specified in the manuscript, patients not requiring supplemental oxygen therapy or transfer to the intensive care unit (ICU) were classified as mild/moderate (N = 166), while those that did were classified as severe/critical (N = 58).

### Estimating the distribution of seroconversion times

One of the goals of this study is to estimate the means and variation of IgG and IgM seroconversion times (time between symptom onset and first antibody detection) for different assays, antigens, and disease severity. We developed a custom bootstrapping procedure to do this using data on seroconversion times that have been reported in a diverse number of ways (from exact days to periods up to 22 days, and as raw results for one individual or means for groups of individuals). Our approach ensures that the best data (i.e. high-resolution data in the form of one specific seroconversion time for one individual) have the most influence on estimates of the means and standard deviations (sd) of seroconversion times. The stepwise weighted bootstrapping procedure integrates all types of data that contain useful information about the timing of seroconversion of different antibodies in day(s) post symptom onset (dpo). The first step of the bootstrap used only the highest-resolution data on seroconversion (i.e. reported for exact days, as opposed to a range of days), while subsequent steps included data with decreasing levels of resolution. When seroconversion times were reported for groups of individuals simultaneously, each individual group member was treated as a separate individual that can be sampled randomly. Seroconversion times were sometimes reported as a mean time (± standard error; se) instead of an exact time or time period. In these cases, the standard deviation of time around the mean was calculated (using reported sample size and standard error), and a random time was drawn from this normal distribution. Data from cumulative seroconversion curves were incorporated by assigning the seroconversion time at which the curve increases to the number of individuals being reported to seroconvert at that time. In the bootstrapping procedure, each of these individuals could then be sampled in the same way as any other individual.

At each step, a distribution of observed possible seroconversion times was bootstrapped 50,000 times from repeated random sampling of individual seroconversion times from the dataset. At the end of each bootstrapping step *n*, a normal distribution was fitted to the obtained distribution of possible seroconversion times. This distribution was then used as prior information to weight sampling probability during the weighted bootstrapping procedure at step *n* + 1 (Lee & Young 2003). Additional details are provided in Supplementary Information. Aside from increasing sample size (and hence the confidence in the estimates) and the density of the histogram/distribution, there were no significant differences between distributions estimated using different maximum time periods (Supplementary Information Figure S1).

### Detection probability of IgG and IgM

The probability of detecting SARS-CoV-2 specific IgG or IgM in plasma or serum samples was estimated by integrating data on whether an individual tested positive or negative on a given dpo. Data containing information on detection probability on a given day are reported in diverse ways, using different resolutions of sample size (from one individual to results reported for groups) or time (results reported on specific days or as a range of days). Additionally, time series data from individuals sampled multiple times contain information about detection probability for times between measurements. These diverse data sources were integrated using different rules. When antibody levels were reported, the cut-off provided in the studies was used to determine the negative or positive status of samples. Individual results for a specific day were included as reported. When time was reported as a period, the midpoint time was used. When a proportion of positive samples was reported together with a sample size, the number of positive and negative samples were calculated and used as independent samples. When two samples that are part of a longitudinal time series showed the same result, the individual was assumed to have the same result for all times within the interval. When such samples had different results, the (interpolated) samples in the early half of the interval were assigned the same result as the first sample, and those in the later half were assigned the same result as the second sample. This procedure resulted in a dataset where each dpo has a number of positive and negative observed samples that could be used to estimate a daily detection probability. Binomial exact confidence intervals of the means were added.

### Detection probability of RNA

The probability of detecting RNA in upper and lower respiratory samples, and in faecal samples, was estimated using the same procedure used for IgG and IgM, but excluding the assumption that days in the interval between two samples of a time series have the same result, i.e. not including any interpolated samples. This was based on the fact that RNA shedding has been observed to be highly variable (Kucirka *et al*. 2020; Wölfel *et al*. 2020). Respiratory sample types were classified as upper (saliva, naso- or oropharyngeal) or lower (sputum, tracheal aspirate, bronchoalveolar lavage) respiratory tract samples. As RT-PCR protocols based on different target sequences resulted in similar sensitivities (Sethuraman *et al*. 2020), all data were pooled for our analysis of detection probability.

### Antibody level kinetics

To characterize the kinetics of antibody levels, we fit models to all individuals for whom longitudinal data were available (i.e. at least three samples are available, one of which has to be positive). Our goal was to estimate the rate of increase, and the timing and magnitude of the peak antibody level. Assays, antigens and reporting units differed extensively between studies, so antibody levels were normalized by dividing the level of each sample in a study by the maximum value observed in that study. This allowed us to compare antibody level kinetic patterns between different studies. All time series are shown in Supplementary Information.

As there were no (or very limited) data available for the later phase of kinetics, when antibody levels decay from their peak, we focused on the early phase of antibody increase up to peak level. These early-phase dynamics follow a standard growth rate pattern, for which well-described functions are available. Of these functions, a three-parameter Gompertz function, *y*(*t*) = *ae^−be^−ct^^*, was an excellent candidate, as its three parameters correspond to clinically significant measures of antibody level (*y*) dynamics over time (*t*). The asymptote (*a*) corresponds to the peak level, displacement (*b*) corresponds to the seroconversion time, and growth rate (*c*) corresponds with the titer increase rate. Antibody levels (*y* and *a*) were log-transformed.

We fit this function to the observed time series of normalized antibody levels using Bayesian Markov Chain Monte Carlo inference. Details can be found in Supplementary Information. Briefly, parameters *a* and *b* were fit separately for each individual, and parameter *c* was shared by all individuals (i.e. the same rate for each individual). Posterior means of the parameters were used for further analyses and for plotting, and can be generated using the accompanying R code (Supplementary Information). Data were combined into subsets depending on the measure of interest (assay, targeted antigen, disease severity). Peak antibody level timing was approximated as the time at which the level reaches 95% of the maximum level (*a*).

All data preparation, cleaning, analysis and plotting was done in R version 3.6.1 (R Core Team 2019) using packages ggplot2 (Wickham 2016), dplyr (Wickham *et al*. 2019), readxl (Wickham & Bryan 2019), patchwork (Pedersen 2019), binom (Dorai-Raj 2014), tidyr (Wickham & Henry 2019) and ggridges (Wilke 2020). Welch two-sample t-tests were used to test for differences between estimated distributions. When reporting ELISA results in the main text, IgG results are shown for assays using NP as target antigen (ELISA-NP), and IgM results are shown for assays using the Spike antigen (ELISA-Spike), as these assays are most often used for the two antibody types (Sethuraman *et al*. 2020; To *et al*. 2020).

## Results

### The distribution of seroconversion times

For the estimation of seroconversion times, 270 data points from 99 individuals were used for IgG, and 240 data points from 71 individuals for IgM. Mean IgG seroconversion time is 13.3 days post symptom onset (dpo) when using ELISA-NP, and 12.6 dpo for IgM using ELISA-Spike (Figure 1a). These results do not differ significantly (t = 0.22, df = 7.7, P = 0.84), and are similar when using MCLIA (Figure 1b). Variation in seroconversion times is substantial regardless of assay, and for both IgG (sd = 5.7) and IgM (sd = 5.8).

Disease severity does not significantly affect seroconversion time, for IgM or for IgG ((Figure 1c–d). Mean IgG seroconversion time for mild/moderate cases is 12.9 dpo, vs 15.5 dpo for severe/clinical cases (t = –0.96, df = 14.8, P = 0.35). Mean IgM seroconversion time for mild/moderate cases is 12.3 dpo, vs 13.2 dpo for severe/critical cases (t = –0.2, df = 23.5, P = 0.83). A detailed overview of seroconversion time results including means and standard deviations is provided in Supplementary Information (Figures S2 to S5; Table S1).

**Figure 1.**
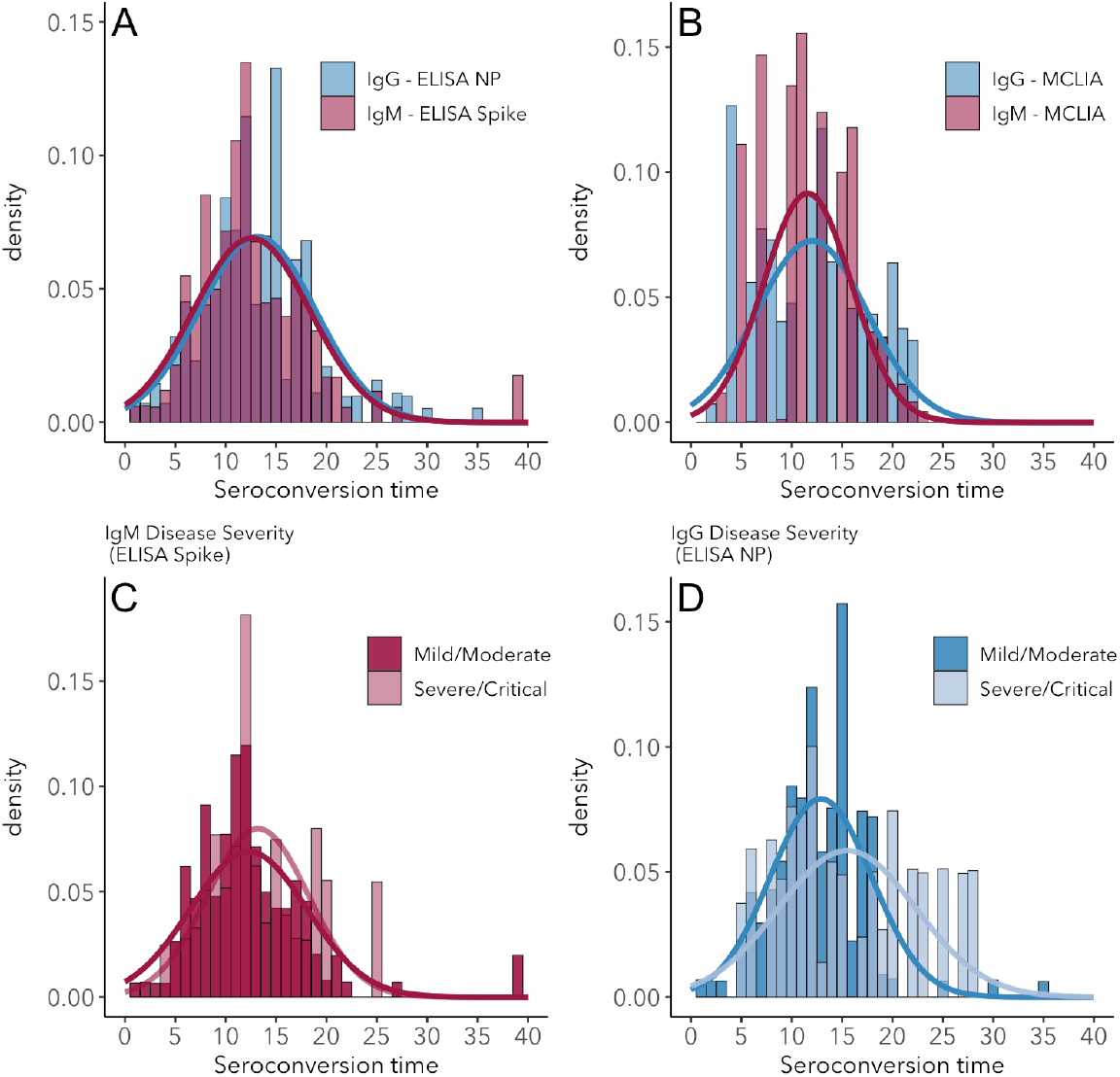
Seroconversion time distributions for IgG and IgM. (A) IgG and IgM detected using ELISA. (B) IgG and IgM detected using MCLIA. (C) IgM and (D) IgG seroconversion related to disease severity. IgG and IgM ELISA results are shown for the NP and Spike antigens, respectively. Times are reported as days post symptom onset.

### Antibody detection probability

While estimates of seroconversion time provide information about the first moment at which antibodies can be detected, changes in detection probability over time provide useful information about the proportion of individuals that has detectable antibodies. Sample sizes for observed and interpolated data that can be used for antibody detection probability (see methods) are 8,053 for IgG and 7,935 for IgM, with daily mean sample sizes of 224 and 220, respectively. The probability of detecting IgG increases over time, reaching a maximum around 25–27 dpo, at which point between 98 and 100% of individuals test positive for the presence of IgG (Figure 2a). Detection probability remains at this maximum level for the remainder of the days available in the studies existing at the time of writing (up to 60 days). This pattern is consistent across assays (Figures S6–7). IgM detection probability is similar to that of IgG until its peak near 90% around 25 dpo, but starts to decrease shortly after, reaching a 50% detection probability around 60 dpo (Figure 2a). Although data on neutralizing antibody presence were sparse, we observe that detection probability rapidly rises to near 100%, where it remains up to the last time available in the dataset (29 dpo). Detection probability patterns do not differ significantly between mild/moderate and severe/clinical cases, aside from a slightly steeper rise for severe/critical cases (Figure S9). Raw antibody detection probability data are provided in Supplementary Information S1.

### RNA detection probability

Samples sizes for observed and interpolated data are 7,443 and 1,793 for upper and lower respiratory samples and 1,179 for faecal samples, with mean daily sample sizes of 226, 72 and 39.3, respectively. The probability of detecting viral RNA in respiratory and faecal samples is high (80–100%) at symptom onset, and is highest for lower respiratory tract samples. Detection probability decreases rapidly at rates dependent on sample type (Figure 2b), and most rapidly for upper respiratory tract samples, but the proportion of positive samples approaches zero around 30 dpo for each sample type. Raw antibody detection probability data are provided in Supplementary Information S1.

**Figure 2.**
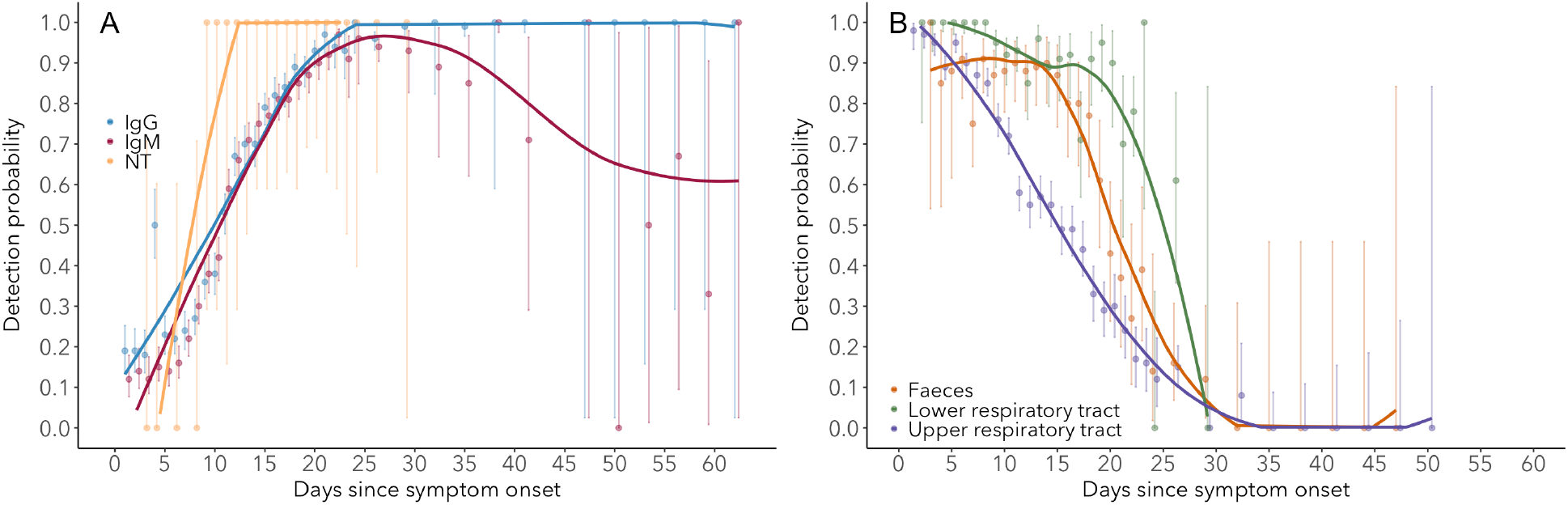
Detection probability of IgG, IgM and NT (neutralizing) antibody (A) and RNA in different sample types (B) over time since symptom onset. Points are mean values for each day. Bold lines are flexible smoothed splines fit the data. Error bars indicate binomial exact 95% confidence intervals of the mean, based on daily sample size. Note that error bars after day 30 tend to be large, due to the limited available data. IgG and IgM values are those detected using ELISA (target antigens NP for IgG and Spike for IgM). After day 25, results are pooled into 3-day periods in order to improve estimates.

### Antibody level kinetics

Patterns of antibody level kinetics are relatively consistent across antibody type, assay and antigen (Figure 3). Although there is strong individual variation in seroconversion timing (as reported above) and peak level, this variation does not correlate significantly with assay or antigen (Figure 3a). Peak level is reached around 16–17 dpo and does not differ significantly between IgG and IgM, measured using ELISA (IgG mean = 16.6±0.5 dpo±se; IgM mean = 16.9±0.6 dpo±se; t = 0.5, df = 39.8, P = 0.61), except for IgG when measured using ELISA Spike (mean = 21.6±0.5 dpo±se; t = 7.3, df = 37.4, P < 0.0001). Disease severity does not significantly affect the time at which peak levels are reached, for IgG (t = 1.9, df = 14.7, P = 0.08) nor IgM (t = 1.1, df = 17, P = 0.3) (Supplementary Information). All assays exhibit comparable growth rates, although IgG ELISA-Spike increases slower than the other assays, and IgG ELISA-NP slightly faster than IgM ELISA-Spike (t = 2.6, df = 25.4, P = 0.01).

**Figure 3.**
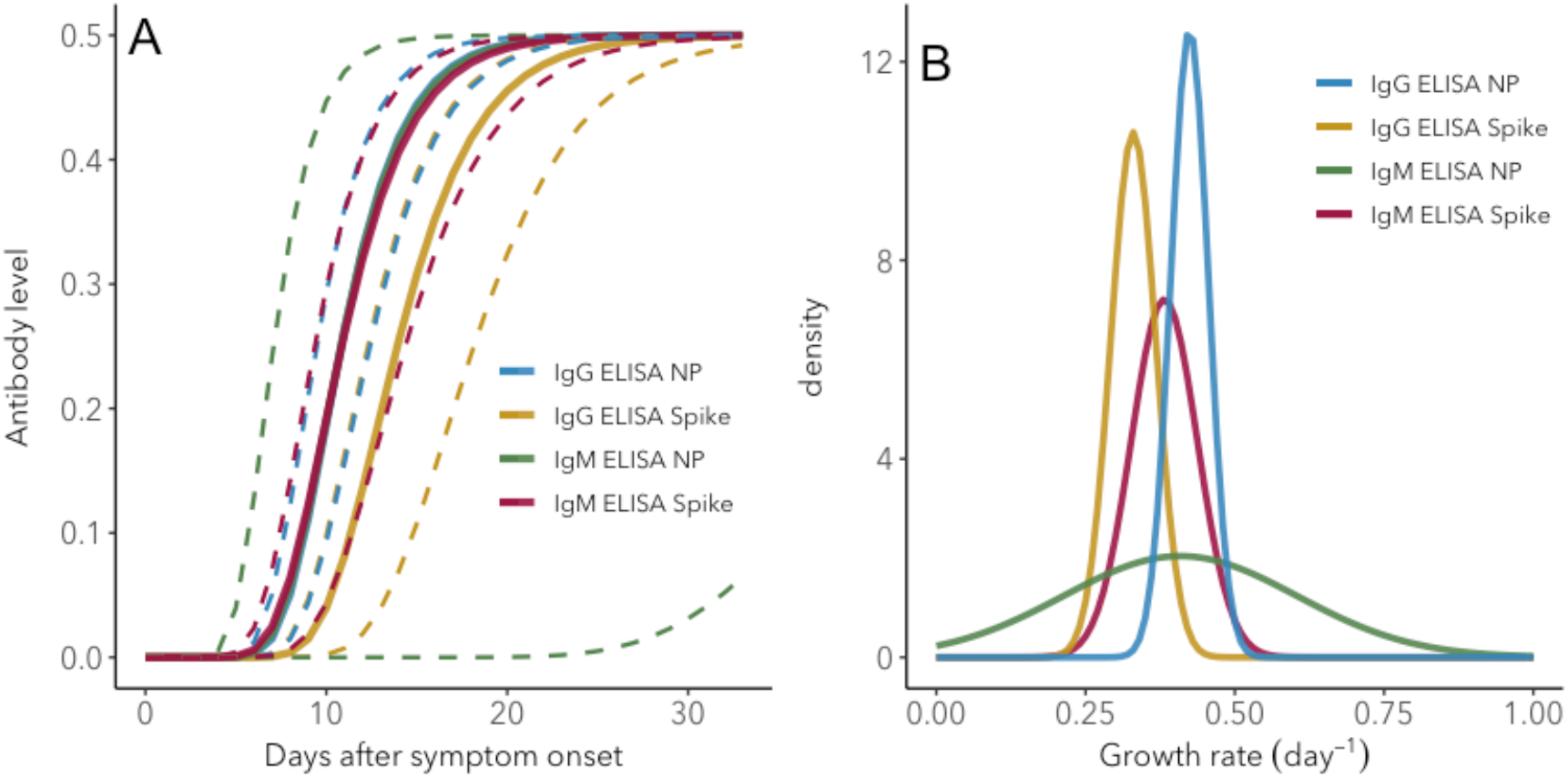
IgG and IgM antibody level kinetics for different antibodies and assays. (A) Fitted functions using posterior mean values for increase rate and start of the increase phase (displacement). Dotted lines show upper and lower 95% credible intervals. Note that the mean functions for all assays except ELISA Spike IgG, as well as the upper interval line for ELISA Spike IgG, overlap (B) Normal distributions fitted to the posterior distributions of the increase rate.

## Discussion

By leveraging multiple data sources on key aspects of the antibody response against SARS-CoV-2, we were able to produce quantitative estimates of the mean and variation of seroconversion timing, antibody level kinetics, and the changes in antibody and RNA detection probability. These results provide critical reference information for serological surveys, transmission models, and herd immunity assessments. By combining data from 22 different studies using different assays, antigens, protocols and patient groups, we were able to quantify the means and, crucially, the extent of variation of important serologic and RNA shedding parameters.

Seroconversion time is highly variable between individuals, with a mean around 12–13 days post symptom onset. We find that IgG and IgM can be detected as early as 0 dpo in 10 –20% of patients, which indicates that seroconversion can happen at, and likely before the onset of detectable symptoms. To our knowledge seroconversion prior to symptom onset has not been observed, which is likely due to the fact that such cases are typically not under investigation. By integrating a wide range of data sources we find more extensive variation in seroconversion timing than previously observed, and importantly it was possible to quantify the distributions around the mean seroconversion times (Haveri *et al*. 2020; Huang *et al*. 2020; Zhao *et al*. 2020). One potential caveat related to any type of result reported as time since symptom onset is that variation in the incubation period (time between infection and symptom onset) can affect the estimated timing of antibody kinetics and RNA shedding. The mean incubation period is estimated to be around 7–8 days, with a standard deviation of 4.4 (Ma *et al*. 2020). The clear antibody and RNA detection patterns we observe here suggest that the effect of this variation does not obscure broad patterns, but if incubation period does differ in certain groups of individuals, relative results may be affected. This could indeed be the case for disease severity, as mild cases are estimated to have a longer incubation period (8.3 days) than severe cases (6.5 days) (Ma *et al*. 2020).

Patterns of IgM and IgG detection align with immunological expectations, as IgM antibodies are typically present during the early phase of the immune response while IgG antibodies remain detectable for much longer periods (Xiao *et al*. 2020). We detected IgG and IgM antibodies in nearly all (98–100%) individuals by day 22–23 after symptom onset, consistent with recent findings (Kraay *et al*. 2020). While IgG detection remains at this level for at least the remainder of available times in the dataset (60 days), the proportion of IgM-positive samples decreases again, reaching 60–70% by 60 dpo. In other words, the proportion of individuals losing IgM increases from day 30 onwards. The quantification of changes in detection probability over time is relevant for clinical testing, as this relates to test sensitivity (Sethuraman *et al*. 2020).

It has been postulated that disease severity and humoral immunity against SARS-CoV-2 are correlated, but results so far have been inconclusive (Okba *et al*. 2020). Here, we did not detect any significant effects of disease severity on antibody patterns. For IgM, this matches findings from a 28-case cohort study, where seroconversion appeared to be the same for severe and non-severe cases (Tan *et al*. 2020). In that same study however, IgG data on 45 cases suggested that seroconversion may be earlier for severe cases, and it was hypothesized that disease severity may affect antibody response characteristics. Similarly, earlier seroconversion in severe cases has been observed for SARS-CoV-1 (Lee *et al*. 2006), but this result was not consistent across studies (Chan *et al*. 2005). The consensus patterns from our larger integrative study suggest that if disease severity is indeed linked to antibody response, the effect is subtle and sensitive to other sources of variation, explaining the inconsistencies seen across studies. In addition, if disease severity indeed does not significantly affect antibody patterns and protective immunity develops even in mild cases, these cases would be substantial contributors to herd immunity development. This finding may also be important for vaccine efficacy, however it is not yet known whether IgG or IgM development correlates with protective immunity (Altmann *et al*. 2020), although we do observe that neutralizing antibodies appear at an even higher rate than IgG or IgM (Figure 2a).

The extensive amount of individual variation in antibody patterns, which is a common phenomenon (Pacis *et al*. 2014) may affect the accuracy of transmission models (Weitz *et al*. 2020). For example, if seroconversion times reflect the actual onset of immunity (i.e. protection against reinfection), the observed range of 0 to 40 dpo may need to be included in the transition rate parameter from the exposed to the infectious class in SEIR-type models (Li *et al*. 2020). When conducting a study using such data, it is therefore important to carefully consider how this variation may affect results, and whether it should be taken into account explicitly (Wearing *et al*. 2005), especially given the potentially enormous impact of COVID-19 transmission models (Kissler *et al*. 2020).

We observed clear patterns of RNA shedding that have several important implications, particularly for sampling designs. First, it is clear that the probability of detecting RNA is highly dependent on sample type, consistent with previous observations (Memish *et al*. 2014; Tan *et al*. 2020). Lower respiratory tract samples have the highest probability of testing positive for SARS-CoV-2 RNA, particularly after about 15 dpo. During the first 8 dpo, 100% of samples tested positive for RNA. While detection probability for faecal and upper respiratory tract samples are nearly as high at symptom onset as lower respiratory tract samples, it decreases much more rapidly, with the lowest average detection probability for upper respiratory samples. Nevertheless, it appears that by 30 dpo detection probability approaches zero for all sample types, although it is important to note that the dataset did not include lower respiratory samples beyond day 29, which means that the true shedding endpoint in lower respiratory samples could not be determined. These results match those from multiple studies (Guo *et al*. 2020; Sethuraman *et al*. 2020; Tan *et al*. 2020; To *et al*. 2020). When interpreting results on RNA shedding, it is important to note that the presence of RNA does not necessary imply the presence of live virus (Theel *et al*. 2020; Wölfel *et al*. 2020).

Together, these antibody and RNA detection probability patterns provide an excellent reference for informing sampling designs. Figure 4 provides an overview of the key patterns. Based on the information of interest, different designs would be optimal:

**Figure 4.**
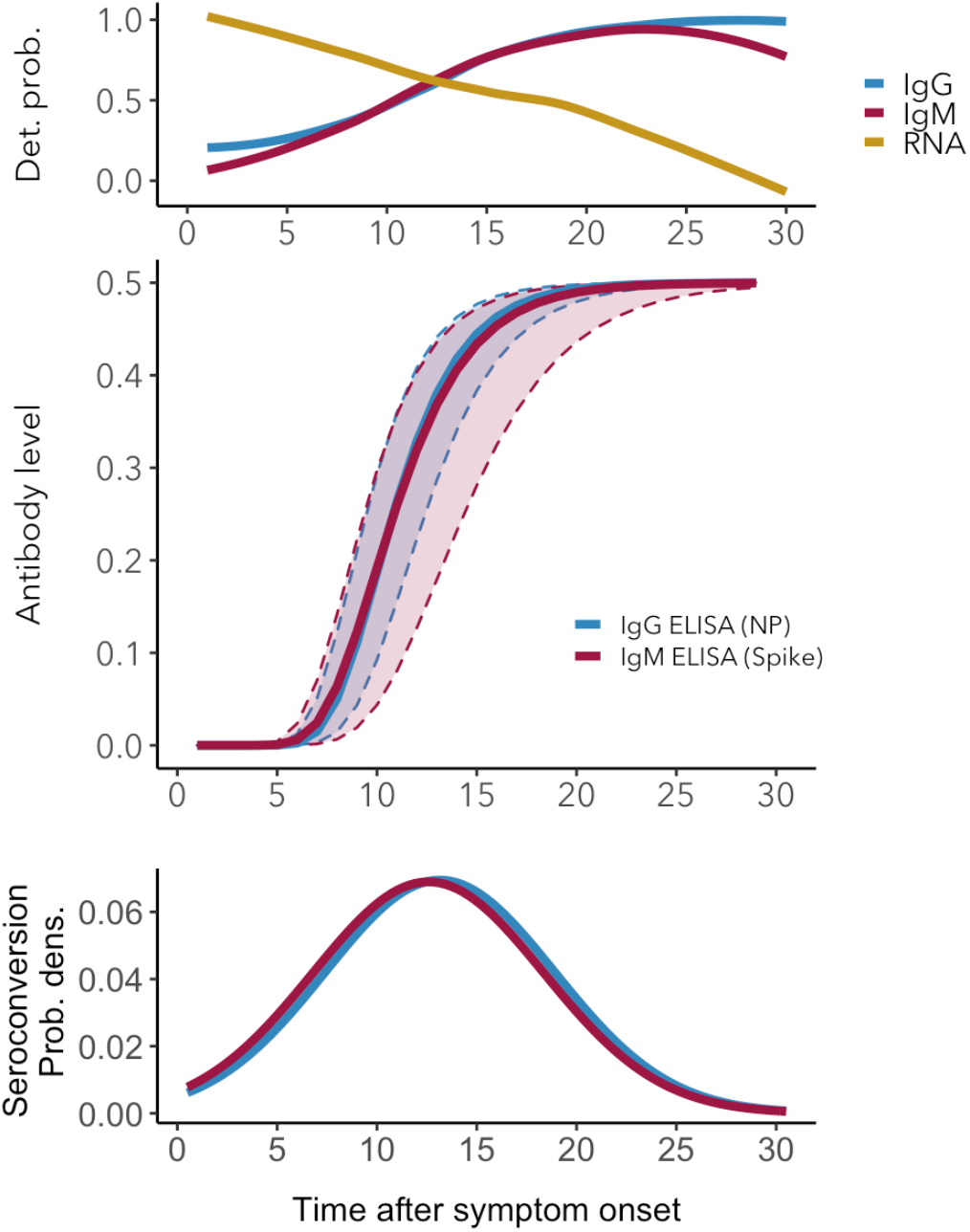
Antibody and RNA shedding patterns during the early phase of SARS-CoV-2 infection. (Top) Detection probability of IgG, IgM and RNA in upper respiratory tract samples. (Middle) Mean IgG and IgM level kinetics with 95% credible intervals. (Bottom) Distribution of observed IgG and IgM seroconversion times.

- To determine whether an individual has been exposed in the past: test for the presence of IgG after 25 dpo, and at least up to day 60 (and possibly much longer, up to 1 or 2 years, depending on how long IgG antibodies persist; Chang *et al*. 2005). Particularly important for transmission models (Kucharski *et al*. 2020; Weitz *et al*. 2020) and herd immunity (Lassaunière *et al*. 2020; Theel *et al*. 2020).
- To determine the ratio between IgG and IgM as a measure of recent vs older exposure: test for IgG and IgM between 20 and 30 dpo. Particularly important for distinguishing recent from older infection (Chang *et al*. 2005; Du *et al*. 2020) and contact tracing, notably for asymptomatic cases (Okba *et al*. 2020).
- To determine whether an individual is shedding: test for the presence of viral RNA as soon as possible after symptom onset, and preferably using multiple samples taken on different days due to variability (Wölfel *et al*. 2020). Consider trade-offs between detection probability in different sample types and invasiveness when sampling. An IgM-positive test also indicates recent infection and might be used as a parallel test. Particularly important for assessing transmission risk to others and contact tracing (Giordano *et al*. 2020).

In summary, this study provides a reference of key antibody and RNA shedding parameters, including estimates of variation that can be used to inform serological surveys and transmission models. As research on SARS-CoV-2 is highly active and more data will undoubtedly become available, parameters can be updated through the use of the algorithms made available in the accompanying R code.

## Data Availability

Data are available as Supplementary Information

## Acknowledgments

BB was supported by the European Commission Horizon 2020 Marie Sklodowska-Curie Actions (grant no. 707840). JOL-S and AG are supported by the Defense Advanced Research Projects Agency DARPA PREEMPT # D18AC00031 and the UCLA AIDS Institute and Charity Treks, and JOL-S and KCP are supported by the U.S. National Science Foundation (DEB-1557022), the Strategic Environmental Research and Development Program (SERDP, RC‐2635) of the U.S. Department of Defense and the Cooperative Ecosystem Studies Unit Cooperative Agreement #W9132T1920006.

## Data availability

The compiled dataset is available as Supplementary Information S2. The R markdown file used for data preparation, analysis and visualization is available as Supplementary Information S3.

## Footnotes

1. https://www.cdc.gov/coronavirus/2019-ncov/hcp/clinical-guidance-management-patients.html

